# Specialty Visits and Real-world Initiation of Cardioprotective Anti-hyperglycemic Medications Among US Adults with Type 2 Diabetes

**DOI:** 10.1101/2022.05.17.22275232

**Authors:** Arash A Nargesi, Callahan Clark, Mengni Liu, Abraham Reddy, Samuel Amodeo, Rohan Khera

## Abstract

Prescription of sodium glucose cotransporter-2 inhibitors (SGLT-2i) and glucagon-like peptide-1 receptor agonists (GLP-1RA) among patients with guideline-directed indications remains limited with substantial inter-prescriber variability. In this prospective study of US adults, we used administrative claims database of individuals with type 2 diabetes and compelling indications for SGLT-2i and GLP-1RA to evaluate the impact of healthcare visits with certain specialty providers on the initiation of these medications. These specialties included family medicine, internal medicine, cardiology, endocrinology, and nephrology. Overall, 294,988 individuals eligible for SGLT-2i and 198,525 for GLP-1RA were identified. In 2019-2020, SGLT-2i and GLP-1RA were initiated in 10.4% and 16.7% of eligible individuals, respectively. After accounting for patient characteristics and comorbidities, healthcare visit with endocrinologists was associated with the highest rate of initiation of either drug across specialties (OR=2.16 [2.08-2.24] for SGLT-2i, and 2.76 [2.64-2.88] for GLP-1RA). Healthcare visits with cardiologists and with family medicine and internal medicine physicians were only modestly associated with initiation of SGLT-2i and GLP-1RA. The study highlights the need for broad education for expansion of the use of these medications rather than focus on dedicated specialty clinics.

## BACKGROUND

Clinical practice guidelines recommend sodium glucose cotransporter-2 inhibitors (SGLT-2i) and glucagon-like peptide-1 receptor agonists (GLP-1RA) for individuals with type 2 diabetes (T2D) and high cardiovascular risk. Fewer than 1 in 10 patients with guideline-directed class I indications receive these medications in the US.^1^ With substantial prescriber-level variation for SGLT-2i and GLP-1RA prescriptions,^2^ the impact of healthcare visits with specific specialty providers on the use of these medications has not been well characterized. In a prospective nationwide cohort of US adults with compelling indications for SGLT-2i and GLP-1RA use followed for 2 years, we evaluated whether care received from certain clinical specialists portended successful initiation of these essential drugs.

## METHODS

We used Optum Lab’s de-identified administrative claims data from beneficiaries with Medicare Advantage Part D or commercial insurance representing a mixture of ages and geographical regions from across the US. Individuals 18 years of age or older with T2D and American Diabetes Association and American Heart Association guideline-based comorbidity indication for SGLT-2i or GLP-1RA with 36 months of continuous enrollment from 2018-2020 were included. To ascertain new initiators, the study identified individuals without pharmacy claims for these medications throughout 2018. These individuals were followed from January 2019 through December 2020, with pharmacy claims defining initiation of treatment.

Comorbidity-based indications included atherosclerotic cardiovascular disease (ASCVD), heart failure, and diabetic nephropathy for SGLT-2i and ASCVD for GLP-1RA.^3^ Exclusion criteria were chronic kidney disease stage IV-V, end stage renal disease, and dialysis for SGLT-2i; and medullary thyroid carcinoma and multiple endocrine neoplasia syndrome type-2 for GLP-1RA.

The study exposure was one or more visits with clinicians of specific specialties that provide care to patients with T2D. These specialties included family medicine, internal medicine, cardiology, nephrology, and endocrinology. Study outcome was defined as new initiation of SGLT-2i and GLP-1RA.

In multivariable logistic regression, we evaluated whether receiving care from a specific clinician group was associated with higher initiation of cardioprotective agents. We adjusted for factors that may portend patient-level differences, including age, sex, neighborhood income, insurance type (Medicare vs Commercial), Charlson comorbidity index (CCI), and frequency of clinical encounters parametrized as tertiles as a surrogate for healthcare needs. For SGLT-2i, indication of use was included in the model. Data were analyzed using R (CRAN-3.4.0) with 2-tailed p<0.05 to define significance.

## RESULTS

We identified 294,988 individuals eligible for SGLT-2i and 198,525 for GLP-1RA. Among those eligible for SGLT-2i, 62.5% had at least one encounter with family medicine, 62.0% with internal medicine, 57.2% with cardiology, 21.0% with nephrology, and 14.4% with endocrinology. Among those eligible for GLP-1RA, 61.9% had at least one encounter with family medicine, 62.9% with internal medicine, 64.4% with cardiology, 16.6% with nephrology, and 13.7% with endocrinology (**Table**). Overall, 30,707 individuals with an indication for SGLT-2i (10.4%) and 33,311 for GLP-1RA (16.7%) initiated the medication in 2019-2020.

**Table.** Baseline characteristics of individuals with type 2 diabetes and guideline-based indication for SGLT-2i and GLP-1RA. Data are presented as mean (SD) for age, median [IQR] for income and frequency of clinical encounters, and number (%) for others. Frequency of clinical encounters include total number of outpatient visits per year. Abbreviations: SGLT-2i, sodium glucose cotransporter-2 inhibitor; GLP-1RA, glucagon-like peptide-1 receptor agonist; CCI, Charlson comorbidity index; ASCVD, atherosclerotic cardiovascular disease.

|  | **SGLT-2i** | **GLP-1RA** |
| --- | --- | --- |
| **Number** | 294,988 | 198,525 |
| **Age (year)** | 75.5 (8.6) | 75.8 (8.3) |
| **Female** | 147,131 (49.9%) | 91,426 (46.1%) |
| **Income (\$)** | 52,784 [46,338-62,149] | 52,782 [46,338-62,036] |
| **Insurance** |  |  |
| **Medicare** | 175,135 (93.3%) | 185,316 (93.3%) |
| **Commercial** | 19,853 (6.7%) | 13,209 (6.7%) |
| **CCI Score** | 5.1 (2.4) | 5.1 (2.6) |
| **ASCVD** | 202,335 (68.6%) | 198,525 (100%) |
| **Heart Failure** | 66,959 (22.7%) | 45,736 (23%) |
| **Diabetic Nephropathy** | 155,270 (52.6%) | 70,149 (35.3%) |
| **Total Annual Clinical Encounter** | 13 [8-21] | 13 [8-21] |
| **Specialty Encounter** |  |  |
| **Cardiology** | 168,811 (57.2%) | 128,014 (64.5) |
| **Nephrology** | 62,058 (21%) | 33,044 (16.6%) |
| **Endocrinology** | 42,490 (14.4%) | 27,278 (13.7%) |
| **Family Medicine** | 184,500 (62.5%) | 122,899 (61.9%) |
| **Internal Medicine** | 183,015 (62%) | 124,867 (62.9%) |

In multivariable models adjusted for demographic and comorbidity profiles, healthcare visit with endocrinology was associated with the highest rates of initiation of either drug across specialties (OR=2.16 [2.08-2.24] for SGLT-2i, and 2.76 [2.64-2.88] for GLP-1RA) after accounting for differences in patient characteristics. Healthcare visits with cardiologists and with family medicine and internal medicine physicians were only modestly associated with initiation of SGLT-2i and GLP-1RA (**Figure)**. Diabetic nephropathy and nephrology visits were associated with lower initiation rates for SGLT-2i (OR=0.94 [0.90-0.99] and 0.7 [0.67-0.74], respectively), but not GLP-1RA.

**Figure.**
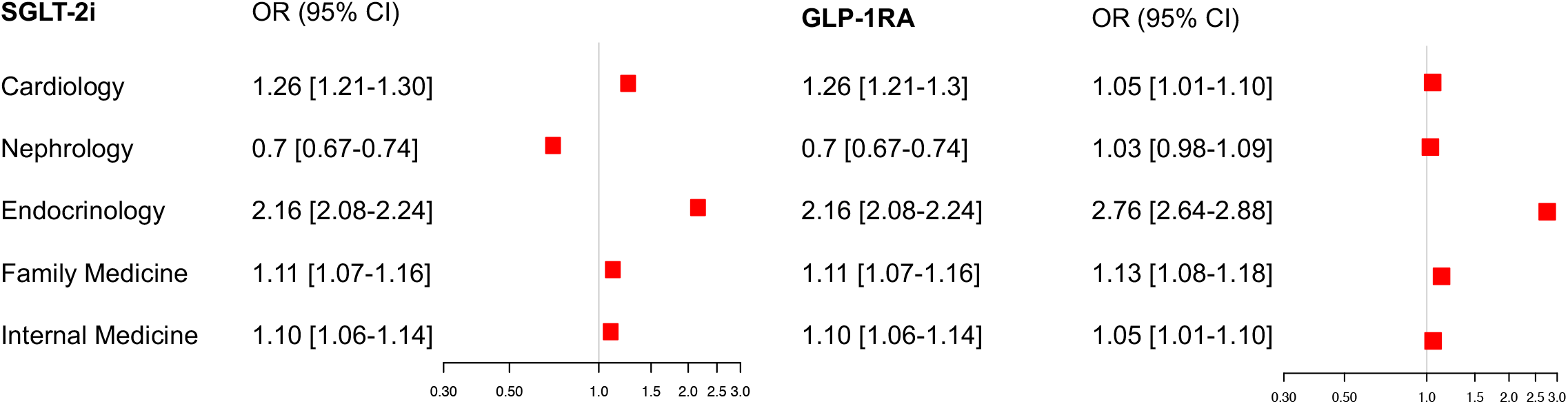
Multivariable predictors of SGLT-2i and GLP-1RA initiation. Data represent odds ratio for each specialty visit adjustment for age, sex, income, insurance type, Charlson comorbidity index, frequency of clinical visits, other specialty visits, and in case of SGLT-2i indication of use. Abbreviations: SGLT-2i, sodium glucose cotransporter-2 inhibitor; GLP-1RA, glucagon-like peptide-1 receptor agonist.

## DISCUSSION

Individuals receiving care from endocrinologists have 2-to 3-fold higher rates of SGLT-2i and GLP-1RA initiation. In contrast, initiation rates among those receiving care from cardiologists were comparable to those from family medicine and internal medicine providers, and those with nephrologists were substantially lower. The study does not suggest a uniform benefit from specialty vs. general medical care, and highlights a need for broad education, dissemination, and uptake rather than an approach that focuses on dedicated specialty clinics.

The strength of the study is the nationwide assessment of patients across age groups. The limitation is a lack of assessment of glycemic control and identifying the exact provider who prescribed the medication for patients visiting more than one specialty clinician. However, the study represents individuals with high cardiovascular risk, all of whom have an indication for these agents, regardless of their glycemic control.

## Data Availability

The data are proprietary and are not available for open sharing due to restrictions on our data use agreement

## DATA AVAILABILITY

The data are proprietary and not available for open sharing due to restrictions on our data use agreement.

